# Validating Artificial Intelligence Guidance for Ultrasound Acquisition and Remote Interpretation

**DOI:** 10.64898/2026.07.16.26356882

**Authors:** Thomas Maldonado, Satish Muluk, Parth Rali, Nilam Soni, Robert Nathanson, Hani Kuttab, Matthew VandeHei, Collin Michels, John Swietlik, Giancarlo Speranza, Olivia Schaffer, collaborating investigators group, Fouad Al Noor, Sven Mischkewitz, Bernhard Kainz, Mike Blaivas, Glenn Jacobowitz

## Abstract

Diagnostic pathways for deep vein thrombosis (DVT) requiring ultrasound are limited by availability and prolonged time-to-imaging. This multicenter, double-blinded, prospective, nonrandomized study evaluated the performance of an AI-guidance system (ThinkSono Guidance, ThinkSono, GmbH) enabling non-ultrasound-trained operators to acquire proximal lower extremity compression ultrasounds for remote clinician interpretation. Patients underwent AI-guided ultrasound(s) and imaging specialist-performed ultrasound(s). Primary and secondary endpoints were image quality, sensitivity/specificity for proximal DVT, and prioritization specificity (effectiveness in identifying patients requiring further ultrasound after AI-guided scan). Of 634 recruited subjects, 594 subjects (700 scans) with 67 DVTs were analyzed. 86.83% of AI-guided scans achieved diagnostic image quality; triage sensitivity was 92.86%, triage specificity 39.12%, prioritization specificity 97.96%. Imaging specialist-performed ultrasounds could be avoided in 35.32% of patients. Median AI-guided scan and review time was 7.57 minutes. These findings suggest AI-guided, clinician-reviewed ultrasound may be a scalable triage strategy to expand DVT evaluation access, particularly in resource-constrained and after-hours settings.

## Introduction

Venous thromboembolism (VTE) constitutes a major global health problem with an annual incidence of over 10 million cases, roughly 150 cases per 100,000 individuals.^1^ The clinical sequelae of VTE are severe, ranging from life-threatening pulmonary emboli (PE) to debilitating post-thrombotic syndrome, contributing to significant long-term morbidity, mortality, and healthcare expenditures.^2^

The standard diagnostic approach for evaluating suspected lower extremity deep vein thrombosis (DVT) often relies on ultrasound, typically performed by a sonographer (often duplex ultrasound) or a trained clinician at the bedside (compression point-of-care ultrasound, POCUS).^3-7^ Hereafter, both are referred to as specialist ultrasound, to differentiate from ultrasound performed by staff without extensive ultrasound training.

Access to timely ultrasound is often limited by the availability of qualified personnel and requisite equipment. Some surveys have shown only a quarter of major academic medical centers maintained on-site ultrasound technicians during evening and overnight hours, and recent data from two major U.S. medical centers suggest most VTE diagnoses occurred more than 72 hours after initial presentation.^3,8^ Over 80% of patients investigated for DVT will be negative, expending significant resources, leading to ultrasound backlogs which could delay positive diagnoses.

Any diagnostic delay could carry significant consequences. Untreated DVT can progress to pulmonary embolism in approximately 50% of cases, with PE mortality rates of 10-30% when undiagnosed.^1,2,9^ Even when DVT is ultimately diagnosed, prolonged time-to-treatment increases the risk of clot propagation, post-thrombotic syndrome, and recurrent VTE.^1,2,9^ The 72-hour diagnostic window reported by Kang et al. represents a period during which patients remain at substantial risk for preventable complications.^3^

One approach to address these limitations is expanding ultrasound access with an AI-based guidance system allowing non-ultrasound trained operators to acquire ultrasound images. Real-time anatomic and technique guidance is provided to operators without ultrasound training, allowing for acquisition of venous compression ultrasound images. These images are transmitted to qualified clinicians for rapid remote interpretation (**Figure 1**). One such technology has been studied in clinical trials across the United Kingdom and Europe, leading to its adoption into clinical practice (**Figure 2**).^10-15^ The objective of this pivotal study is to evaluate the clinical performance of this AI-guided compression ultrasound acquisition system within the United States healthcare environment.

**Figure 1.**
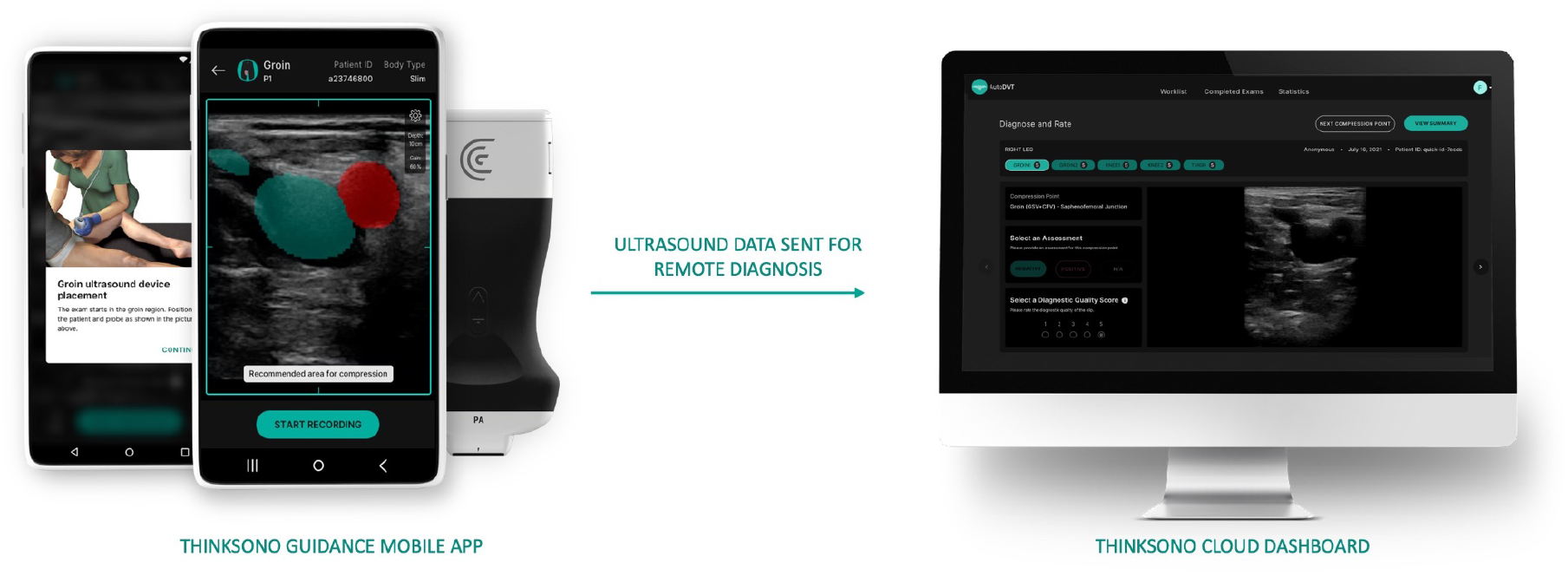
AI guided ultrasound workflow and system overview. Overview of the AI guidance platform architecture, including the mobile acquisition interface, cloud-based clinician review dashboard, and integrated end-to-end clinical workflow.

**Figure 2.**
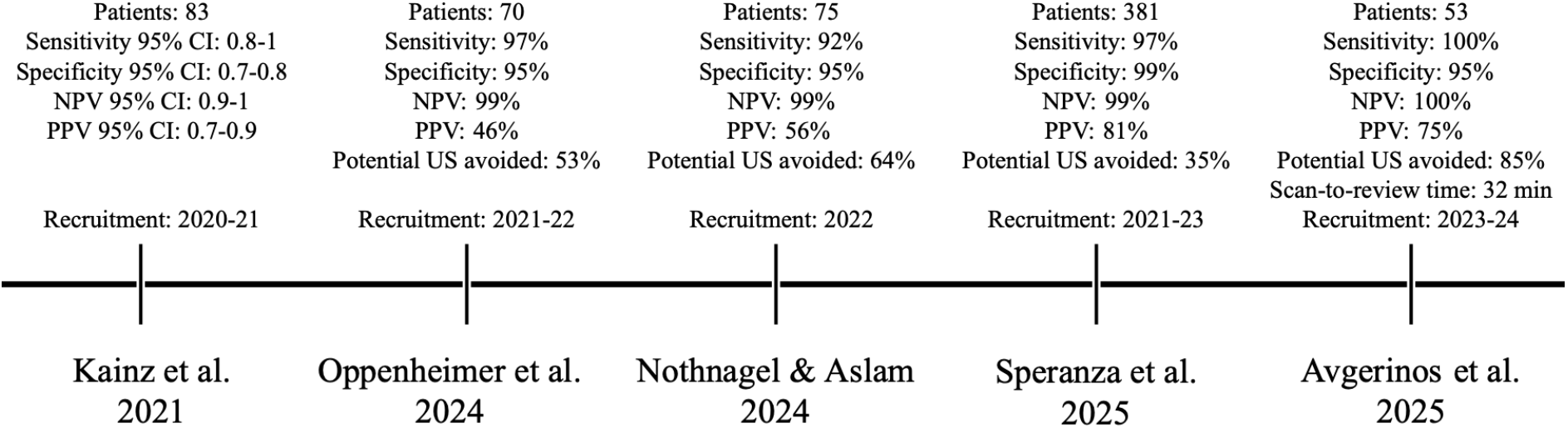
Timeline of clinical investigations. Clinical investigations evaluating AI-guided compression ultrasound triage sensitivity, prioritization specificity, and other endpoints compared to standard care. (CI, confidence interval; NPV, negative predictive value; PPV, positive predictive value; US, ultrasound)

## Methods

“Determining the Validity of ThinkSono Guidance for Ultrasound Image Acquisition and Remote Detection” (DVT GUARD) is a prospective, multicenter, double-blinded study conducted across five sites. The study is registered on www.clinicaltrials.gov (NCT06652568, submitted Oct 12, 2024). All sites are tertiary or quaternary care institutions of varying sizes across different regions of the United States. Sites followed the same study protocol, with site-specific recruitment targets. Institutional review board ethics approvals were obtained at each site, and written informed consent was obtained from all patients by onsite research team members trained in human subjects research ethics.

### Study design

Patients presenting with clinical indications for lower extremity venous ultrasound were eligible for enrollment. Subjects in this study underwent an AI-guided compression POCUS scan conducted by operators without prior ultrasound training. Operators included clinical research personnel (n=7), physicians in training (n=5), clinical staff (nurses, medical assistants, and patient care technicians) (n=6), non-physician providers (n=1), and medical and allied professions students (n=5) across participating sites. None were credentialed or certified to perform venous compression ultrasound, though some had exposure to ultrasound use and non-diagnostic point-of-care applications (e.g., IV placement guidance). Subjects also underwent local (i.e. per the study site institution) standard of care (SOC) lower extremity specialist-performed ultrasound scans conducted by sonographers and interpreted by local hospital-credentialed clinicians.

All parties remained blinded throughout the study. AI-guided scan operators received no scan results or indication of DVT detection. Operators did not communicate with the remote qualified clinicians (RQCs) or local imaging specialists. RQCs are blinded to the local imaging specialist ultrasound findings and to each other’s evaluations and do not communicate with operators or other RQCs. Local imaging specialists and interpreting clinicians are blinded to the AI-guided scan results and do not see the AI-guided scans. Patient flow is shown in Supplementary **Figure 1**.

### Inclusion and exclusion criteria

Inclusion criteria were patients willing and able to provide informed consent, above the age of 18, with signs and symptoms suggestive of DVT for whom a lower extremity venous ultrasound is indicated.

Exclusion criteria were patients unwilling or unable to give consent, patients undergoing a local imaging scan and initiation of new treatment for before an AI-guided scan can be performed, patients without a local imaging specialist scan, patients with incomplete AI-guided scans, and pregnant patients.

### AI guidance system description

The AI-guided compression ultrasound system (ThinkSono Guidance, ThinkSono GmbH, Potsdam, Germany) is a guidance, data acquisition, and communication tool (**Figure 1**). This system is based on a machine learning vessel segmentation network; this convolutional neural network classifies the anatomical landmark plane in the current field-of-view, scores vein compressibility, and provides semantic segmentation masks for arteries and veins. It is trained to identify blood vessels, anatomic landmarks, and vessel compression states. It has been validated in multiple pre-clinical and clinical studies.^10-15^ ThinkSono Guidance is FDA 510(k) cleared (Number: K260338) and EU MDR Class IIb approved, and is in use across the UK and EU.

Non-ultrasound-trained healthcare professionals receive 60-90 minutes of training in operating the system and following system prompts and guidance on probe placement/movement, target anatomy location, and vein compression. Users are not trained to perform a compression ultrasound exam themselves. The AI system performs quality control analysis on the images. The AI guidance system does not generate any diagnostic determination. The images are transmitted to RQCs for review and interpretation. Exams can be assessed as:

- Compressible: All cine-loops of adequate quality and all cine-loops show adequate venous compressibility.
- Incompressible: All cine-loops of adequate quality and at least one cine-loop shows venous incompressibility.
- Indeterminate: At least one cine-loop of inadequate quality or at least one cine-loop with indeterminate compressibility due to technique or technical difficulty.

### Outcomes

The primary endpoint was at least 60% of AI-guided scans achieve adequate image quality (a score of ≥ 3 on the American College of Emergency Physicians (ACEP) image quality scale, Supplementary Table 1) as rated by RQCs. This threshold was selected based on prior physician-performed POCUS studies reporting adequate image quality rates of 70-90% when performed by trained clinicians, and literature suggesting that a guidance system approaching this benchmark would meaningfully impact clinical care, workflows, and volume management.^10,12,16-18^ The 60% threshold was considered the minimum acceptable performance for regulatory and clinical viability.

The number of confirmed DVTs is defined as the number of patients for which the local reference scan reports proximal lower extremity DVT. This local reference scan is considered a “ground truth” scan (i.e. the definitive clinical result used for comparison). Thus, only confirmed proximal lower extremity thromboses are included as “positive for proximal DVT” ground truth. Otherwise, the ground truth was considered “negative for proximal DVT.”

Secondary endpoints included measures of discriminative ability based on expert review of AI-guided scan cine-loops. Sensitivity (also referred to as triage sensitivity) and specificity (also referred to as triage specificity) of RQC compressibility assessments were measured and compared to SOC results. Triage sensitivity and specificity were calculated according to **Equations 1 and 2**, respectively (Supplementary **Figure 2)**.

A high triage specificity indicates that downstream imaging specialist resources will be effectively utilized to confirm proximal DVT. An additional specificity metric has been computed to reflect how patients can be prioritized for downstream imaging if normal compressibility cannot be observed. Prioritization specificity considers only ’incompressible’ assessments as test-positive, quantifying the system’s ability to identify high-likelihood DVT cases. A high prioritization specificity indicates that very few false-positives are incorrectly recommended for prioritization. In other words, patients assessed as ‘incompressible’ are indeed more likely to actually have proximal lower extremity DVTs than patients assessed as ‘indeterminate.’ This allows users to prioritize these patients for imaging-specialist evaluation.

Additional variables of interest are included below. Formulas for these endpoint are included in Supplementary Figure 2.

- Potential imaging specialist ultrasounds avoided.
- Triage positive predictive value (PPV) and prioritization PPV
- Time for the operator to perform an AI-guided scan and time for the RQC to review a scan.

### Statistical analysis

Categorical variables are presented as proportions, normally distributed continuous variables are presented as mean ± standard deviation, and skewed continuous variables are presented as median. Statistical significance was determined using a two-sided alpha level of 0.05.

For individual U.S. sites and pooled results, endpoint mean estimates and 95% CIs were computed using nonparametric bootstrapping with resampling at the patient level. Bootstrapping is a well-established and rigorously studied statistical technique for estimating means and confidence intervals, especially in settings where multiple assessments per scan are provided by a panel of reviewers.^13^

For the primary endpoint, the proportion of image adequacy for interpretation as rated by the remote qualified clinicians (RQCs) was computed via bootstrapping by randomly sampling patients and reviewers with replacement for 1,500 iterations. An explanation of the use of bootstrapping can be found in prior publications.^12^ Majority voting analysis across reviewers per patient was also performed. Incomplete AI-guided scan acquisitions were not considered for reporting the proportion of adequate image quality, as this endpoint cannot be reliably measured on incomplete data.

For the secondary endpoint, the sensitivity and specificity of RQCs’ compressibility assessments were computed against the ground truth established by the local imaging specialist standard of care scans. Incomplete AI-guided scans (e.g., partial coverage, missing cine-loops) were retained in the analysis for sensitivity and specificity of triage performance. This reflects the intended clinical workflow of the system, where inadequate imaging defaults to a referral of the patient to an imaging specialist ultrasound scan.

## Results

Recruitment ran from November 2023 to December 2025. Subject flow is displayed in **Figure 3**. Across all 5 sites, 634 patients were recruited, with 594 analyzed and 40 excluded. Exclusion reasons included undergoing treatment between imaging specialist and AI-guided scan (n=7), administrative or logistic issues (e.g. ineligible consent, incomplete enrollment, n=17), aborted scans due to patient limitations (e.g. scan area obstructed by cast, n=13), technical error (n=2), and erroneous lower extremity scan in place of upper extremity scan ordered (n=1). Exclusion reasons are displayed in Figure 3. There were 67 confirmed DVTs on reference scans. Of 594 patients with 700 AI-guided scans, 56 patients (78 scans) had incomplete AI-guided scans and 9 patients (13 scans) lacked imaging specialist scans. This resulted in 585 patients with 687 AI scans considered for triage performance analysis and 538 patients with 622 AI scans considered for image quality analysis.

**Figure 3.**
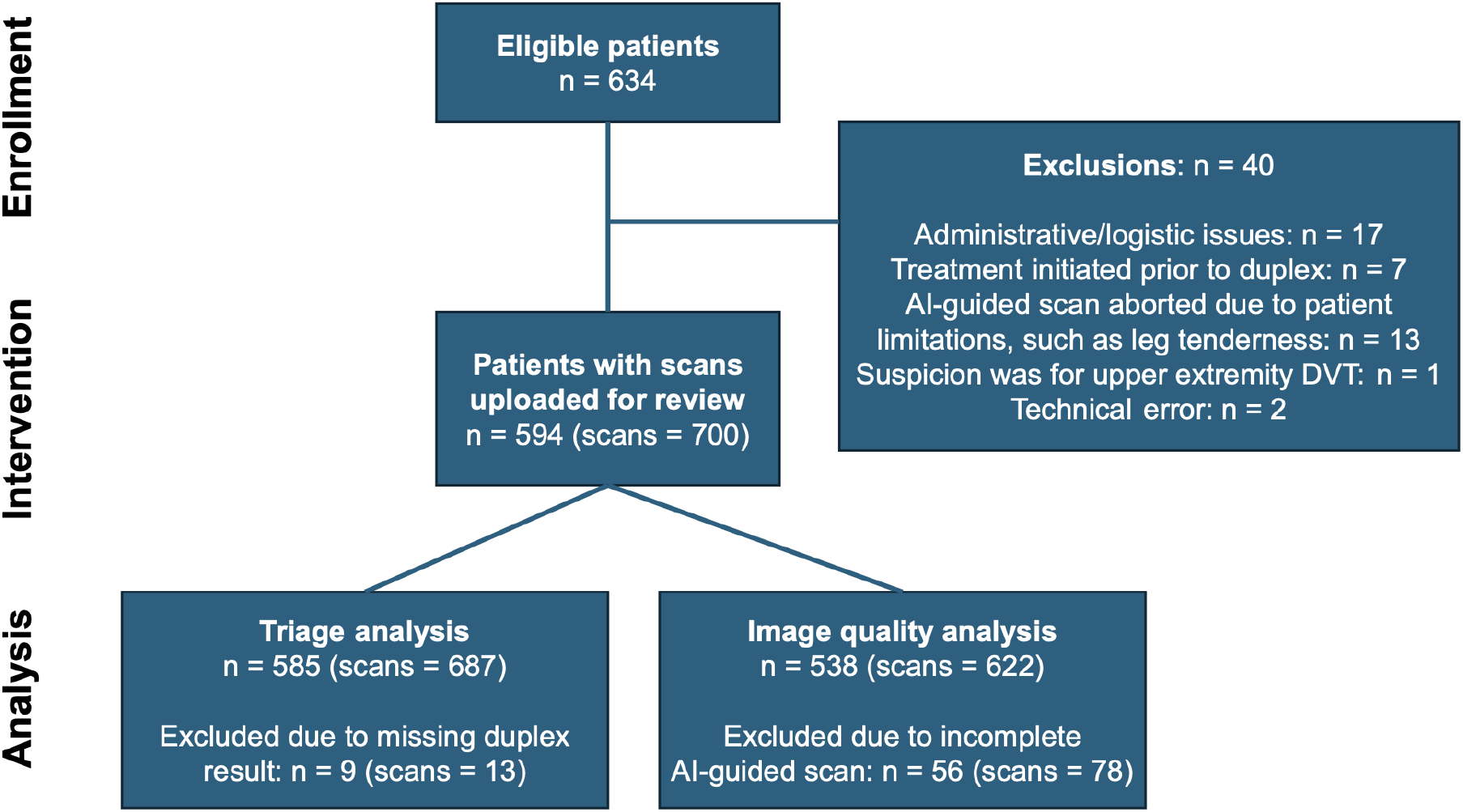
CONSORT Diagram. CONSORT diagram depicting patient flow from enrollment to AI-guided scanning and analysis. Patients are included in the triage analysis if they have imaging-specialist-performed scan results and AI-guided scan results available, regardless of whether the AI-guided scan is complete. Patients are included in image quality analysis if they have complete AI-guided scans, regardless of whether imaging specialist results are available.

Subject characteristics and demographics are displayed in **Table 1**. The overall U.S. cohort was mostly white (61%) and mostly male (60%). The sex distribution of the total cohort was heavily skewed by site E (**Table 2**). Average age was 64 (±15) years and average body mass index (BMI) was 29 (±7) kg/m^2^. Clinical performance data for each site computed via bootstrapping approach are displayed in **Table 2**. Across all U.S. sites, 86.83% (95% CI 84.08-89.34) of scans were adequate for diagnostic interpretation. Sensitivity was 92.86% (95% CI 86.3-98.4) compared to the reference standard. Triage specificity was 39.12% (95% CI 35.18-43.09), and prioritization specificity was 97.96% (95% CI 96.77-99.03). Across U.S. sites, 35.32% (95% CI 31.69-38.99) of imaging specialist ultrasounds could potentially have been avoided.

**Table 1.**
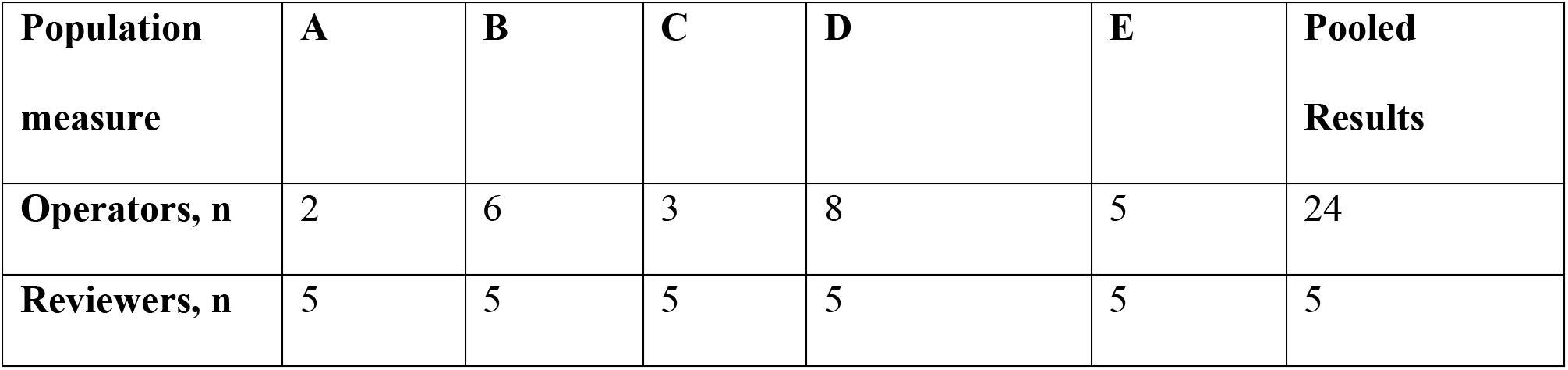
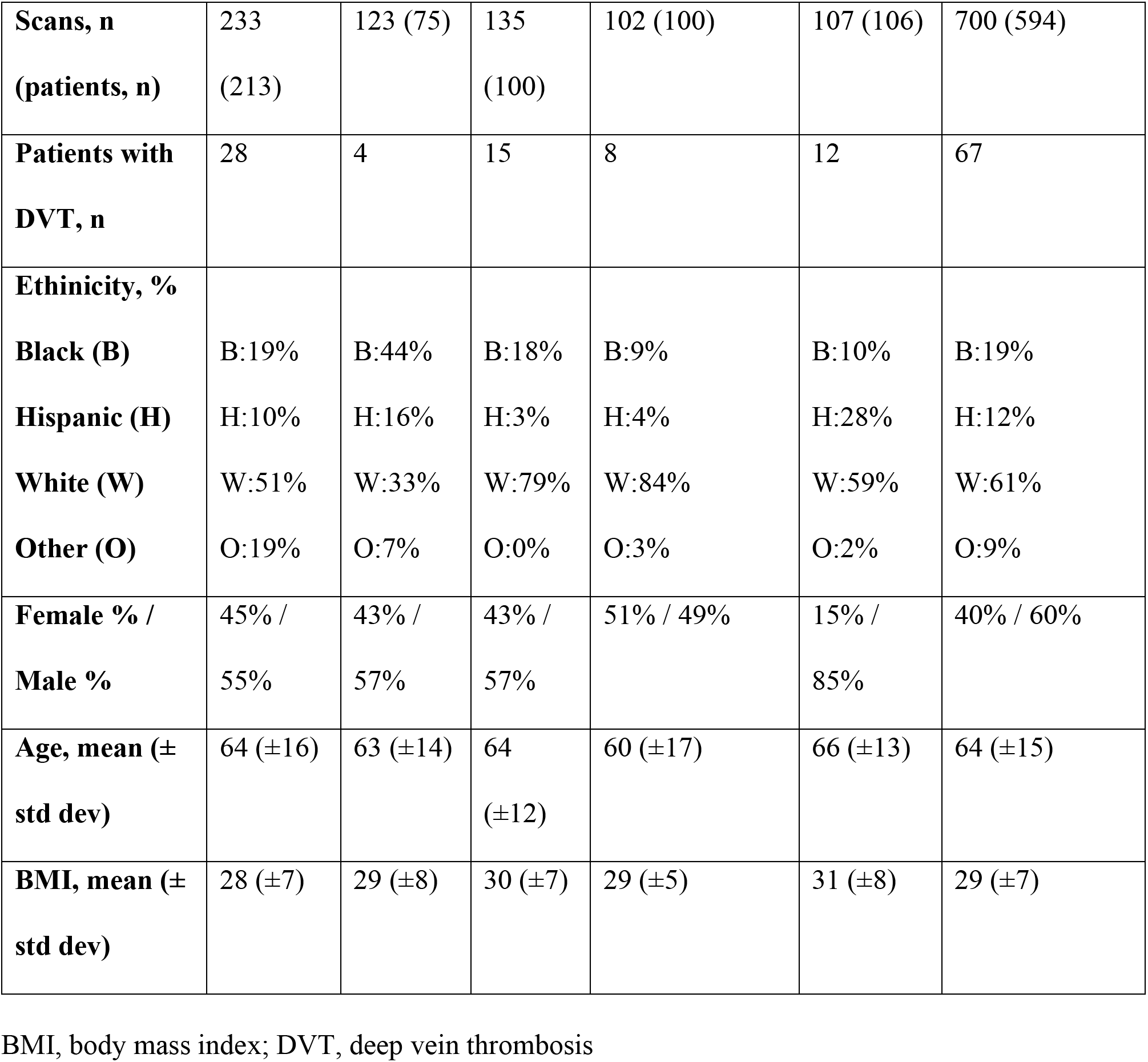
Study population demographics across pooled investigations.

**Table 2.**
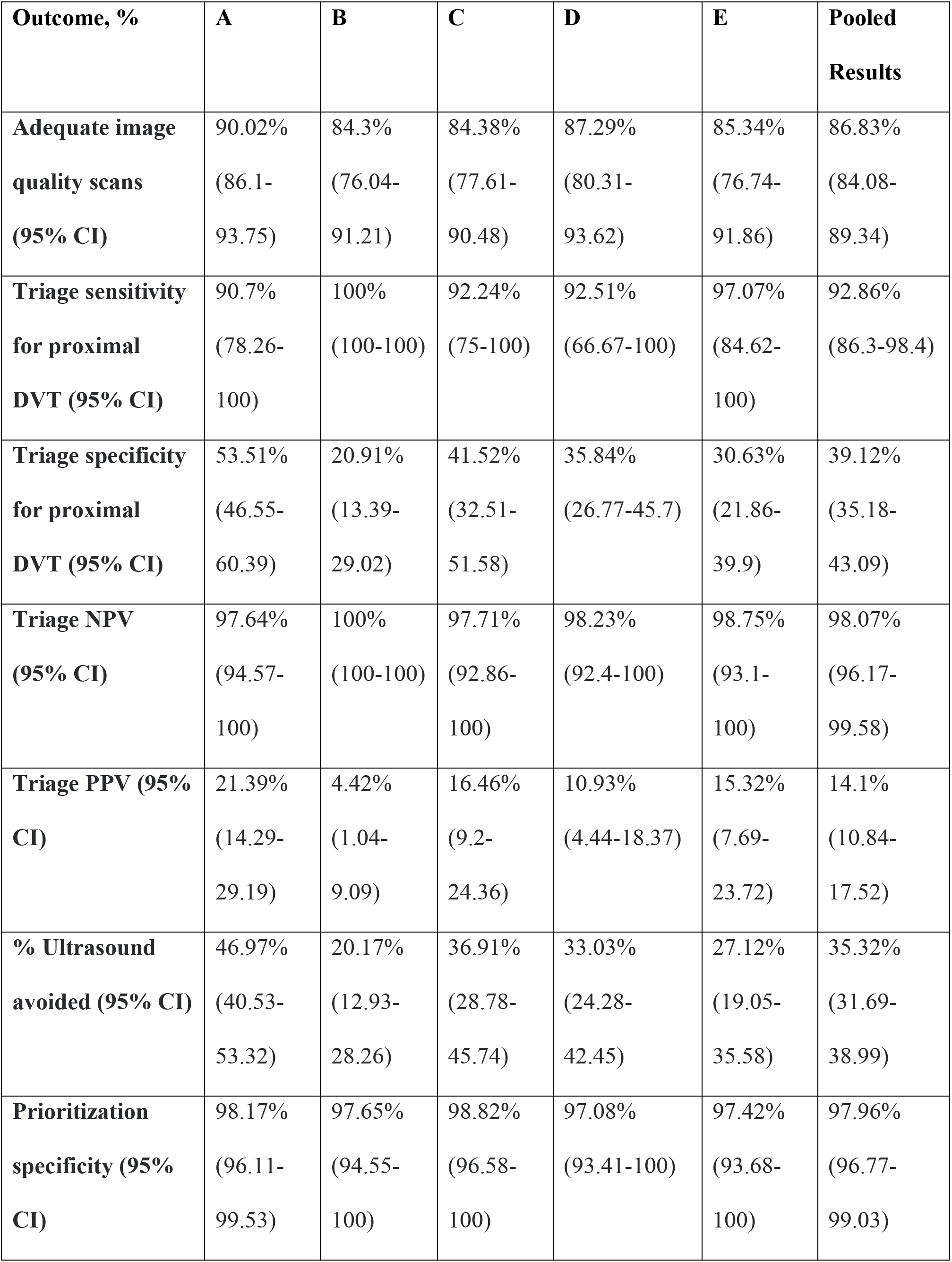
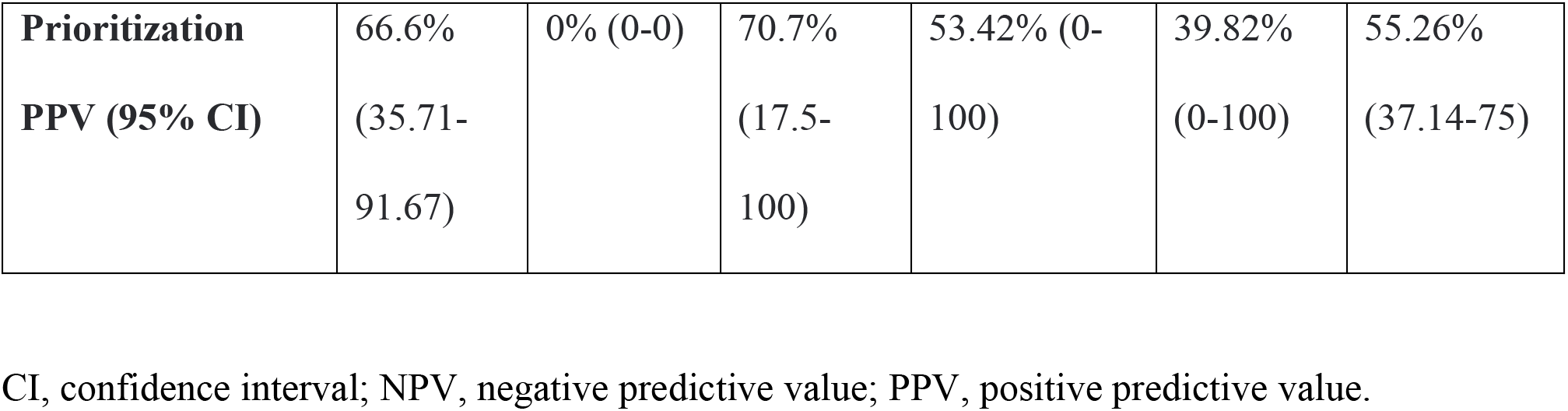
Primary and secondary study endpoints by site and overall, computed by bootstrapping.

Site-level performance varied, with adequate image quality ranging from 84.30% to 90.02% and triage specificity ranging from 20.91% to 53.51%. Notably, sites with higher DVT prevalence tended to show higher triage specificity, consistent with Bayesian expectations that disease prevalence influences predictive value interpretation. **Supplementary Table 2** displays study endpoints by individual RQC and majority voting for pooled U.S. sites. Median time per scan was 5.4 minutes (IQR 5.45 minutes), and median time per review was 2.17 minutes (IQR 2.8 min). There were no adverse events reported in the study.

## Discussion

An AI guidance system enabled non-ultrasound-trained operators to acquire diagnostic-quality images in over 85% of scans with only brief training, supporting feasibility of clinical deployment. Clinicians credentialed to interpret ultrasound demonstrated high sensitivity with strong triage and prioritization performance when reviewing AI guidance-enabled scans, suggesting these qualified clinicians can safely detect proximal DVT while reliably flagging high-risk scans for expedited imaging specialist evaluation. The observed ability to safely avoid imaging specialist ultrasound in over one-third of cases suggests the potential for this system to reduce downstream resource utilization.

Several trends emerged in results across study sites. Though BMI and age were similar across all sites, other demographic features, the prevalence of DVT, and the number of operators differed at each site. The average number of scans per operator at each site also varied, ranging from over 100 at site A to just under 13 at site D. The prevalence of DVT also ranged from 5.3% to 15.0%. Though the studied system produced a high proportion of adequate quality scans and high sensitivity across all sites, differences were noted in triage specificity, triage PPV, and the percentage of ultrasounds avoided (notably at sites B and E). These variables trend together given their formulas (Supplementary Figure 2).

Though the reviewers in this study were consistent across sites, prior work has suggested that different reviewer training and background can impact system performance.^12^ Similarly, the observed differences in outcome measures could be related to differences in operator characteristics, like number of operators or average scans per operator, and/or DVT prevalence. These findings suggest areas for future study to better understand the factors that influence system performance.

Rapid assessment of suspected lower extremity DVT remains a challenge, particularly in emergency and urgent care settings where access to vascular ultrasound expertise is limited. High patient censuses and emergency department staffing constraints, especially outside weekday daytime hours, contribute to diagnostic delays.^19^ In a survey of 79 U.S. teaching hospitals, only 24% had ultrasound technicians on site overnight.^8^ Kang et al. reported that most VTE events in two major U.S. institutions were diagnosed more than 72 hours after presentation, with delayed diagnosis associated with increased 30-day all-cause mortality.^3^ Many facilities lack imaging specialists entirely; only a third of critical access hospitals (CAHs), which serve remote communities and provide care for nearly 20% of the U.S. population, offer ultrasound services.^20-23^

Point-of-care ultrasound (POCUS) has shown promise in addressing these challenges. POCUS has demonstrated good diagnostic accuracy when performed by trained users, with reduced time to diagnosis and reduced ED referrals from general practitioners.^24-27^ However, widespread adoption has been limited by physician shortages, training demands, quality assurance challenges, and lack of physician comfort performing POCUS exams, particularly in resource-constrained settings.^28-35^

AI guidance systems offer one approach to overcome these challenges by allowing for implementation of standard of care POCUS techniques without extensive POCUS training. The AI guidance system studied here has been assessed in multiple trials across the globe with comparable test performance endpoints (Figure 2).^10-15^ The system has demonstrated high sensitivities and specificities, with a significant number of imaging-specialist ultrasounds potentially avoided.

The triage specificity of 39.12% observed in this study represents a 39% potential reduction in imaging specialist ultrasound utilization in DVT-negative patients. From this figure, we see that the AI guidance system enables safe avoidance of imaging specialist ultrasound in over 35% of the total patient population scanned with AI guidance. A 35% reduction in resource utilization is significant; the current diagnostic algorithm for DVT relying on the Wells score and d-dimer testing is valued in part because it estimated to have reduced unnecessary imaging by roughly a third from prior practice.^1,2^ The ability to reduce specialist ultrasound utilization by a further 35% suggests another potentially significant impact on care delivery and efficiency.

Higher imaging specialist ultrasound avoidance rates in prior studies may reflect the impact of real-time communication between operator and reviewer, which was disabled in this study due to the blinding protocol. Avgerinos et al. studied the system with this communication in place, demonstrating an ultrasound avoidance of up to 85% with feedback from remote reviewers with 100% sensitivity when compared against imaging specialist-performed ultrasound.^13^ Disabling communication between operator and reviewer in this meant that scans reviewed as indeterminate remained so, whereas in real-world settings reviewers can request immediate, targeted reacquisition, for example of an incompletely imaged popliteal segment, while the patient is still present. This rapid communication allowing re-acquisition of images to improve quality and aid in reviewer interpretation can positively impact system performance. Collectively, this suggests that system performance without operator-reviewer feedback may underestimate real-world clinical performance.

This suggestion is borne out by evidence from real-world use of the system. Since the conclusion of this study, the system has been deployed in real-world settings with two-way communication enabled. At one urgent care site in the UK National Health Service, 120 patients underwent AI-guided scans over a three month period. The proportion of patients receiving a scan result within four hours of presentation increased from 17.6% prior to system implementation to 51% after implementation. Mean time from presentation to diagnosis fell from 19.6 to 8.9 hours. The number of specialist-performed ultrasounds was reduced by 80.1%. Median time from AI-guided scan initiation to completion of scan review by clinician, including any reviewer-operator communication and re-scanning, was 29 minutes.^36^

Several limitations exist for this study, such as exclusion of certain populations (e.g., pregnant women and those under 18), absence of longitudinal follow-up outcomes, and the fact that the studied AI guidance system did not cover distal (calf) DVT scans at the time of the study. Ultrasound presents significant limitations in detecting distal DVTs, and there are also significant practice and guideline variations regarding scanning and subsequent treatment of distal DVTs.^14,35,37-41^ Certain clinical guidelines recommend a second proximal DVT scan be performed 1 week after the first scan to assess for proximal extension of any distal DVT.^42^ All patients in this study received duplex ultrasound, thus repeat scans were not indicated. This limited the study’s ability to assess the AI guidance system’s sensitivity when used according to clinical guidelines.

We did not assess learning curves at the operator level or assess how individual operator performance may change over sequential scans. Though training on the system does not include training operators to independently perform a compression ultrasound exam, operators still become familiar with the fundamental principles of this examination. This study did not include a control group of to distinguish the effects of the AI guidance system from the impact of a brief training on compression ultrasound on image acquisition. This study demonstrated rapid scan and review times, but did not account for time required for patient preparation, device setup, or image upload time. It also did not assess the feasibility of integration into existing DVT diagnostic algorithms (incorporating d-dimer, Wells score, and clinical decision rules), or cost-effectiveness and economic impacts.

## Conclusion

AI-guided compression ultrasound enables non-ultrasound-trained operators to acquire diagnostic-quality images in 87% of cases, with remote clinician review achieving 93% sensitivity and 98% prioritization specificity for proximal DVT. These performance characteristics are encouraging and support potential clinical applications where imaging specialist ultrasound is unavailable or where ultrasound resources are limited. Over one-third of imaging specialist ultrasounds could be safely avoided, suggesting meaningful potential for resource optimization. Future research should evaluate real-world implementation, cost-effectiveness, and performance in underrepresented populations.

## Data Sharing

Qualified scientific and medical researchers can address requests for de-identified participant data that underlie the results reported in this article to. Proposals for data will be evaluated and approved by ThinkSono, Ltd. at its sole discretion. All approved researchers must sign a data access agreement before accessing the data. Data will be available as soon as possible but no later than within 1 year of the acceptance of the Article for publication and for 3 years after Article publication. ThinkSono, Ltd. will not share data from identified participants or a data dictionary.

## Acknowledgements

The study team would like to thank the numerous sonographers, radiology staff, emergency department staff, and research coordinators that made this study possible. The team would also like to thank all subjects who generously participated in this study.

## Author Contributions

FAN, SM, and GJ conceived the study. FAN, SM, GS, and GJ designed the study and FAN, SM, GS, OS, GJ, TM, PR, SM, NS, RN, HK, MV, CM, and JS helped implement and execute the study across all sites. GS, LA, GB, MB, LC-O, JD, CD, CD, RD, SH, MTH, HI, BJ, AJ, KL, JM, CM, MM, AM, KM, RN, LO, NPK, ZR, TS, and AW recruited and scanned patients. GS and OS conducted a literature review. SM and BK led and supervised statistical analysis. FAN, SM, BK, GS, and OS were responsible for data curation, formal analysis, and data visualization. GS and OS wrote the original draft, which was then reviewed, edited, and approved by all co-authors. All authors had access to the raw data. FAN, SM, TM, and BK verified the data. All authors accept responsibility to submit for publication.

