## Supplement for "Validating Artificial Intelligence Guidance for Ultrasound Acquisition and Remote Interpretation"

### Supplementary Appendix

Table of Contents

| | |
|---|---|
| List of collaborating investigators | 1 |
| Supplementary figures and tables | 2 |
| References | 7 |

List of collaborating investigators

Lizette Aviles[1], Gina Bernardez[2], Matthew Bittner[3], Luis Cardenas-Osorio[3], Jonathan Dempsey[4], Christie Diveterano[3], Christine Doherty[1], Rogelio Duque[1], Suchada Hill[1], Maica Thi Ho[5], Helena Ikenberry[5], Beth Johnson[5], Alexandra Juarez[1], Ka Lio[3], Jennie Matthews[5], Chelsea McClellan[5], Moira Mcgevna[2], Anugya Mittal[3], Kaitlynn Motley[3], Lex Oliver[5], Natasha Pedone-Kahle[5], Zairra Rhodes[4], Taylor Stiegel[4], Audrey Wood[5]

[1]Division of Hospital Medicine, South Texas Veteran's Health Care System, San Antonio, TX
[2]Division of Vascular and Endovascular Surgery, NYU Langone Health, New York NY
[3]Division of Thoracic Surgery and Medicine, Temple University Health, Philadelphia, PA
[4]AHN Cardiovascular Institute, Allegheny Health Network, Pittsburgh, PA
[5]BerbeeWalsh Department of Emergency Medicine. University of Wisconsin-Madison. Madison, WI, USA

Supplementary Figures and Tables

**Supplementary Figure 1. Study Design**

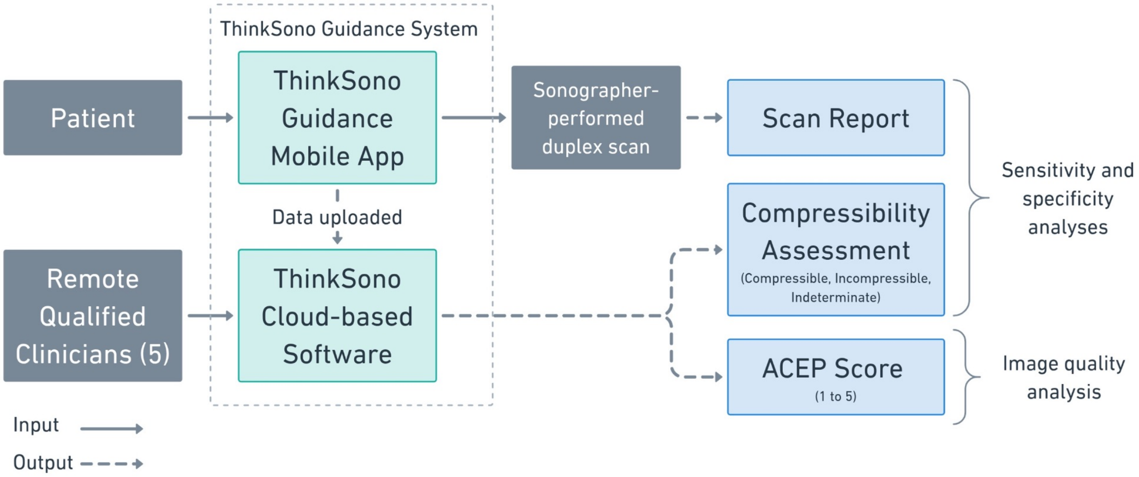


Patients are recruited prospectively and are first scanned by an operator using the AI guidance system. The data is then sent to a remote qualified clinician (RQC) for review. Subsequently the patient is scanned by a local imaging specialist for a final diagnosis. The RQCs are blinded to the results provided by the imaging specialist. All RQCs are also blinded to each other. The operators are also blinded to the imaging specialist result.

**Supplementary Figure 2. Equations**

**Equation 1.** $$sensitivity_{triage} = \frac{\#\, true\ positive_{triage}}{\#\, true\ positive_{triage} + \#\, false\ negative_{triage}}$$

**Equation 2.** $$specificity_{triage} = \frac{\#\, true\ negative_{triage}}{\#\, true\ negative_{triage} + \#\, false\ positive_{triage}}$$

**Equation 3.** $$specificity_{priority} = \frac{\#\, incompressible + \#\, indeterminate - \#\, false\ positive_{priority}}{\#\, incompressible + \#\, indeterminate}$$

**Equation 4.** $$ultrasound_{avoided} = specificity_{triage} * (1 - prevalence)$$

**Equation 5.** $$PPV_{triage} = \frac{\#\, true\ positive_{triage}}{\#\, incompressible + \#\, indeterminate}$$

**Equation 6.** $$NPV_{triage} = \frac{\#\, true\ negative_{triage}}{\#\, compressible\ assessments}$$

**Equation 7.** $$PPV_{priority} = \frac{\#\, high\ priority\ clots}{\#\, incompressible\ assessments}$$

True positive *triage*, incompressible and indeterminate assessments with positive imaging specialist result; true negative *triage*, compressible assessments with negative imaging specialist result; false positive *triage,* incompressible and indeterminate assessments with negative imaging specialist result; false negative *triage,* compressible assessments with positive imaging specialist result; false positive *priority,* incompressible assessments with negative imaging specialist result; PPV, positive predictive value; NPV, negative predictive value; high priority clots, incompressible assessments with positive imaging specialist result.

**Supplementary Table 1. ACEP Grading Scale for Image Quality Assessment[1]**

| ACEP Score | Grading Scale Definition |
|---|---|
| 1 | No recognizable structures, no objective data can be gathered |
| 2 | Minimally recognizable structures but insufficient for diagnosis |
| 3 | Minimal criteria met for diagnosis, recognizable structures but with some technical or other flaws |
| 4 | Minimal criteria met for diagnosis, all structures imaged well and diagnosis easily |
| 5 | Minimal criteria met for diagnosis, all structures imaged with excellent image quality and diagnosis completely supported. |

ACEP, American College of Emergency Physicians

**Supplementary Table 2. Primary Endpoints by RQC and Majority Voting**

| **Outcome, %** | **RQC 1** | **RQC 2** | **RQC 3** | **RQC 4** | **RQC 5** | **Major-ity Voting** |
|---|---|---|---|---|---|---|
| **Proportion of adequate quality scans (95% CI)** | 84.7% (81.7-87.5) | 88.3% (85.6-90.7) | 99.2% (98.2-99.7) | 79.4% (76.2-82.5) | 83.3% (80.3-86.1) | 84.1% |
| **Triage Sensitivity (95% CI)** | 95.5% (87.5-99.1) | 92.5% (83.4-97.5) | 88.1% (77.8-94.7) | 95.5% (87.5-99.1) | 92.5% (83.4-97.5) | 94.0% |
| **Triage Specificity (95% CI)** | 39.4% (35.2-43.7) | 52.6% (48.2-56.9) | 37.1% (33.0-41.4) | 38.4% (34.2-42.7) | 28.7% (24.9-32.8) | 40.7% |
| **Prioritization Specificity (95% CI)** | 100.0% (99.3-100.0) | 99.4% (98.3-99.9) | 80.7% (77.1-84.0) | 98.3% (96.9-99.3) | 98.3% (96.8-99.3) | 100% |

RQC, remote qualified clinician; CI, confidence interval.
